# Cost-effectiveness of real-world administration of chemotherapy and add-on *Viscum album* L. therapy compared to chemotherapy alone in the treatment of stage IV NSCLC patients

**DOI:** 10.1101/2020.02.04.20020354

**Authors:** Anja Thronicke, Thomas Reinhold, Philipp von Trott, Christian Grah, Burkhard Matthes, Harald Matthes, Friedemann Schad

**Affiliations:** Research Institute Havelhöhe at the Hospital Gemeinschaftskrankenhaus Havelhöhe, 14089 Berlin; Institute of Social Medicine, Epidemiology and Health Economics, Charité – Universitätsmedizin Berlin, corporate member of Freie Universität Berlin, Humboldt-Universität zu Berlin, and Berlin Institute of Health, 10117 Berlin; Interdisciplinary Oncology and Palliative Care, Hospital Gemeinschaftskrankenhaus Havelhöhe, 14089 Berlin; Lung Cancer Centre, Hospital Gemeinschaftskrankenhaus Havelhöhe, 14089 Berlin; Medical Clinic for Gastroenterology, Infectiology and Rheumatology CBF and Institute of Social Medicine, Epidemiology and Health Economics, Charité – Universitätsmedizin Berlin, corporate member of Freie Universität Berlin, Humboldt-Universität zu Berlin, and Berlin Institute of Health, 10117 Berlin

**Keywords:** NSCLC, lung cancer, metastatic, stage IV, chemotherapy, add-on VA, cost-effectiveness, integrative oncology

## Abstract

**Background:** For stage IV lung cancer patients receiving add-on Viscum album L. treatment an improved overall survival (OS) was observed. Only limited information regarding cost-effectiveness (CE) for comparisons between standard of care and standard of care plus add-on VA in stage IV lung cancer treatment is available. The present study assessed the costs and cost-effectiveness of standard of care plus VA (V) compared to standard of care alone (C) for stage IV non-small cell lung cancer (NSCLC) patients treatment in a hospital in Germany.

**Methods:** An observational study was conducted using data from the Network Oncology clinical registry. Patients included had stage IV lung cancer at diagnosis and received C or V treatment in a certified German Cancer Center. Cost and cost-effectiveness analyses (CEA) including the analysis of the incremental cost-effectiveness ratios (ICER) were performed from the hospital’s perspective based on routine data from financial controlling department and observed data on OS. The primary result of the analysis was tested for robustness in a bootstrap-based sensitivity analysis.

**Results:** 118 patients (C: n=86, V: n=32) were included, mean age 63.8 years, proportion of male patients 55.1%. Adjusted hospital’s total mean costs for patients from the C and V group were €16,289 (over an adjusted mean OS time of 13.4 months) and €17,992 (over an adjusted mean OS time of 19.1 months), respectively. The costs per additional OS year gained (ICER) with the V-treatment compared to C were €3,586.

**Conclusion:** Based on our analysis, the assessment of inpatient costs and cost-effectiveness of IO concepts in stage IV lung cancer suggests that the combined use of chemotherapy and VA is clinically effective and was comparably cost-effective to chemotherapy alone in our analysed patient sample. Further randomized and prospective cost-effectiveness studies are mandatory to complement our findings.

## Introduction

Integrative oncology (IO) has been increasingly established and acknowledged for the improvement of health-related quality of life during the last decades in international academic and public cancer centres [1, 2]. IO comprises the use of complementary interventions including mind-body-practices, natural products and/or lifestyle modifications in addition to conventional oncological treatment [3]. Complementary mistletoe extracts (*Viscum album Loranthaceae*, VA) which are applied within IO concepts improve the health-related quality of life (HRQL) of oncological patients [4, 5]. The demand for IO is on the rise world-wide due to the increasing self-awareness of people with cancer and cancer survivors who want to self-manage their own care and needs as an active participant in the treatment [6, 7]. IO concepts aim to be accepted in the future as routinely applied health care elements as they contribute to clinical effectiveness by helping people with cancer to maintain their HRQL, to maintain dosing and to avoid interruption of standard oncological treatment. Thus, costs and cost-effectiveness of IO need to increasingly be analysed to determine which of the effective interventions can be delivered to the patients. It has been found that IO concepts have the potential to deliver cost-effective therapies to oncological patients [8, 9]. Lung cancer ranks first position in cancer-related deaths worldwide [10]. The majority (85%) of lung cancer patients are diagnosed with NSCLC and the prognosis of this tumour type is devastating - at a metastasized stage the five-year survival rate is less than 10 percent. Immune checkpoint inhibitors (ICI) which have dramatically improved the clinical outcome of NSCLC patients are costly; some of them are not cost-effective over chemotherapy for NSCLC [11]. Others are cost-effective when therapy selection on PD-L1 expression is warranted [12]. Therefore, the quest for innovative and cost-effective treatment options is ongoing. As limited information on cost and cost-effectiveness (CEA) of IO concepts including add-on VA is available for lung cancer, the purpose of the present study was the CEA of combinational chemotherapy plus add-on VA compared to chemotherapy alone in stage IV NSCLC patients from a German hospital’s perspective.

## Materials and Methods

### Study design, patients and primary objective

A controlled, non-randomized, observational and monocentric real-world study was conducted and data from an accredited oncological registry, Network Oncology, were analyzed [13]. Patients were included in the analysis who were 18 years or older, who gave written consent, with a histologically proven primary diagnosis of stage IV NSCLC receiving standard of care surviving more than 28 days. Patients were not included when death date or last contact date was not available. Follow-up was performed routinely six months after first diagnosis and annually during the next years. Loss to follow-up was defined as no follow-up visits. The CE analysis took the perspective of the hospital Gemeinschaftskrankenhaus Havelhöhe Berlin (GKHB), at which the patients were treated. The GKHB is an Anthroposophic-integrative working hospital harbouring three German Cancer Society (DKG, Deutsche Krebsgesellschaft) - certified Organ Centres, including a certified lung cancer centre. The study served as a feasibility study for subsequent IO cost-effectiveness studies. The primary objective of this analysis was to evaluate the CE of VA in addition to standard of care compared to standard of care alone in stage IV NSCLC patients from the hospital’s perspective.

### Ethics approval and consent to participate

The NO-study has been approved by the ethics committee of the Medical Association Berlin (Berlin— Ethik-Kommission der Ärztekammer Berlin). The reference number is Eth-27/10. Written informed consent has been obtained from all patients prior study enrolment. The study complies with the principles laid down in the Declaration of Helsinki [14].

### Data collection

Structured queries from patient records were run for lung patients (International Classification of Diseases code: C34) using the German Cancer Society accredited clinical registry Network oncology (NO) [13]. Tumour stage at first diagnosis was defined as the earliest recorded stage within a month of the diagnosis date and was coded according to the Union for International Cancer Control (UICC) stages according to the 7th edition of TNM Classification of Malignant Tumours [15]. Demographic hospital-related data such as diagnosis, histology, pre-treatment and treatment as well as last patient record including date of death, the last documentation of personal contact, date of interdisciplinary tumour board conferences or follow-up data were retrieved from the NO registry. Standard oncological treatments and application of VA extracts were queried with their according start and end dates. VA therapy was defined as lasting equal or more than four weeks. Information on hospital stays, length of each hospitalization and longitudinal stationary cost data (according to the German Institute for Hospital Fee Systems (InEK) reporting principles) were retrieved from the hospital’s cost-accounting data base at the financial controlling department. Cost data included the cost of primary therapy for lung cancer, medication, hospital charges for surgery, anaesthesia, diagnostics, laboratories, professional fees, imaging, normal and intensive care units as well as medical and non-medical infrastructure as incurred. Outpatient costs were not included, because they are not captured routinely in the hospital data.

### Allocation of groups

Stage IV NSCLC patients included in the study were classified into one of the two groups: a) C group— patients received only chemotherapy and no add-on VA therapy and b) V group–patients that received combinational chemotherapy and add-on VA therapy. Guideline oriented chemotherapy and add-on VA were applied as per routine clinical care. Non-randomized allocation to the treatment groups was performed by the physician after elaborate information and patient’s decision on treatment options. Applied VA preparations included abnobaVISCUM®, Helixor® and Iscador® VA extracts and were given subcutaneously according to the SmPC [16-18]. Off-label VA application (intravenous, intratumoural) was performed in individual cases.

### Statistical analyses including survival and cost analyses

All statistical analyses were conducted using the software R, version 3.6.1 (2019-07-05 [42] with the exception of sensitivity analysis which was performed by using MS Excel 2016. Data are presented using descriptive statistics, normally distributed continuous data by the mean and standard deviation (SD) or 95% CI and skewed distributions by the median and 95% confidence interval (CI). Binary and categorical variables were presented as absolute and relative frequencies using counts and percentages. For comparison of continuous variables between groups at baseline the unpaired Student’s t-test for independent samples was used. For comparison of categorical baseline variables chi-square analyses were performed. All tests were performed two-sided. P-values <.05 were considered as significant. In survival and cost analyses the distribution of covariates was determined visually via density distribution, quantile-quantile diagrams and/or via frequency distribution.

For the calculation of survival outcome and hazard risks, Kaplan Meier survival analyses, right-censored time-to-event analyses and multivariate stratified Cox proportional hazard analyses were performed as previously reported [19], utilizing the R-packages ‘survival’ (version 3.1–8), ‘prodlim’ (version 2019.11.13), ggplot2 (version 3.2.1) and ‘survminer’ (version 0.4.6). Prior Cox proportional hazard analysis, verification analyses were performed whether or not proportional hazard assumptions were met. The start date for survival analysis (index date) was the date of first histology at diagnosis of stage IV NSCLC. Patient survival was calculated from index date until the patient’s last record. Mean overall survival in months and stationary costs in Euro were adjusted via co-variance analysis by the following co-variates: age, gender (male/female), BMI (4 levels), histology (3 levels), cancer-directed surgery (yes/no), radiation (yes/no), chemotherapy (yes/no), add-on VA therapy (yes/no), smoker status (3 levels). To address cost-influencing factors, multivariable linear regression analyses were performed.

### Cost-effectiveness analyses

For measuring the cost-effectiveness, data on adjusted mean stationary hospital costs was divided by the adjusted mean OS. As a result, the mean hospital costs per month mean OS were reported. In the case the intervention would show a better effect in terms of OS and would be more expensive than the control, the incremental cost-effectiveness ratio (ICER) was calculated. In our analysis the ICER is defined as the additional patient-related costs per additional year OS of the intervention (here V) compared to a control group (here C) and was calculated through dividing the group cost difference by effectiveness difference translating into the mathematical equation 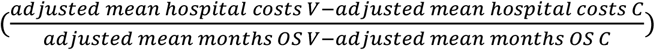. To determine to what extent the primary cost-effectiveness results may vary due to many replications a bootstrap analysis with random 1000-fold resampling population was performed. This analysis accounted for the heterogeneity of individual hospital’s resource consumptions observed in the study. The results of the bootstrap samples were plotted into the four-quadrant diagram (cost-effectiveness plane), which gives graphical information on the incremental results’ robustness.

## Results

### Patients

From 135 patients screened, 17 patients were not included due to missing data, see figure 1. 118 patients (C: n=86, V: n=32) were included for subsequent outcome analysis. Patient’s demographic and clinical characteristics are shown in Tab 1.

**Tab 1:** Baseline characteristics. Characteristics of patients with stage IV non-small cell lung cancer, percentages of sub-characteristics may not add up to 100% due to rounding procedure; n, number of patients; %, percent; SD, standard deviation; C, chemotherapy; V, chemotherapy plus add-on VA; 1) chisquare analysis for categorial variables; Student’s t-test for age distribution

**Tab 1. Baseline characteristics**
|  | All patients (n=118) | C (n=86) | V (n=32) | p-value |
| --- | --- | --- | --- | --- |
| age in years, mean (SD) | 63.8 (10.6) | 63.5 (10.5) | 64.8 (10.8) | 0.58 |
| histology LCC, n (%) | 12 (10.2) | 7 (8.1) | 5 (15.6) |  |
| histology ADC, n (%) | 83 (70.3) | 62 (72.1) | 21 (65.6) |  |
| histology SQC, n (%) | 23 (19.5) | 17 (19.8) | 6 (18.8) | 0.49 |
| gender, female, n (%) | 53 (44.9) | 43 (50.0) | 10 (31.2) |  |
| gender, male, n (%) | 65 (55.1) | 43 (50.0) | 22 (68.8) | 0.11 |
| BMI < 25, n (%) | 56 (47.5) | 40 (46.5) | 16 (50.0) |  |
| BMI 25-29.9, n (%) | 32 (27.1) | 22 (25.6) | 10 (31.2) |  |
| BMI 30+, n (%) | 10 (8.5) | 10 (11.6) | 0 |  |
| BMI NA, n (%) | 20 (17.0) | 14 (16.3) | 6 (18.8) | 0.25 |
| smoker, current/past, n (%) | 89 (75.4) | 67 (77.9) | 22 (68.8) |  |
| smoker, never, n (%) | 9 (7.6) | 8 (9.3) | 1 (3.1) |  |
| smoker, NA, n (%) | 20 (17.0) | 11 (12.8) | 9 (28.1) | 0.10 |
| cancer-directed surgery, yes, n (%) | 10 (8.5) | 6 (7.0) | 4 (12.5) | 0.60 |
| radiation, yes, n (%) | 56 (47.5) | 39 (45.3) | 17 (53.1) | 0.11 |
| comorbidities, n (%) | 50 (42.4) | 36 (41.9) | 14 (43.8) | 0.85 |

**Fig 1:**
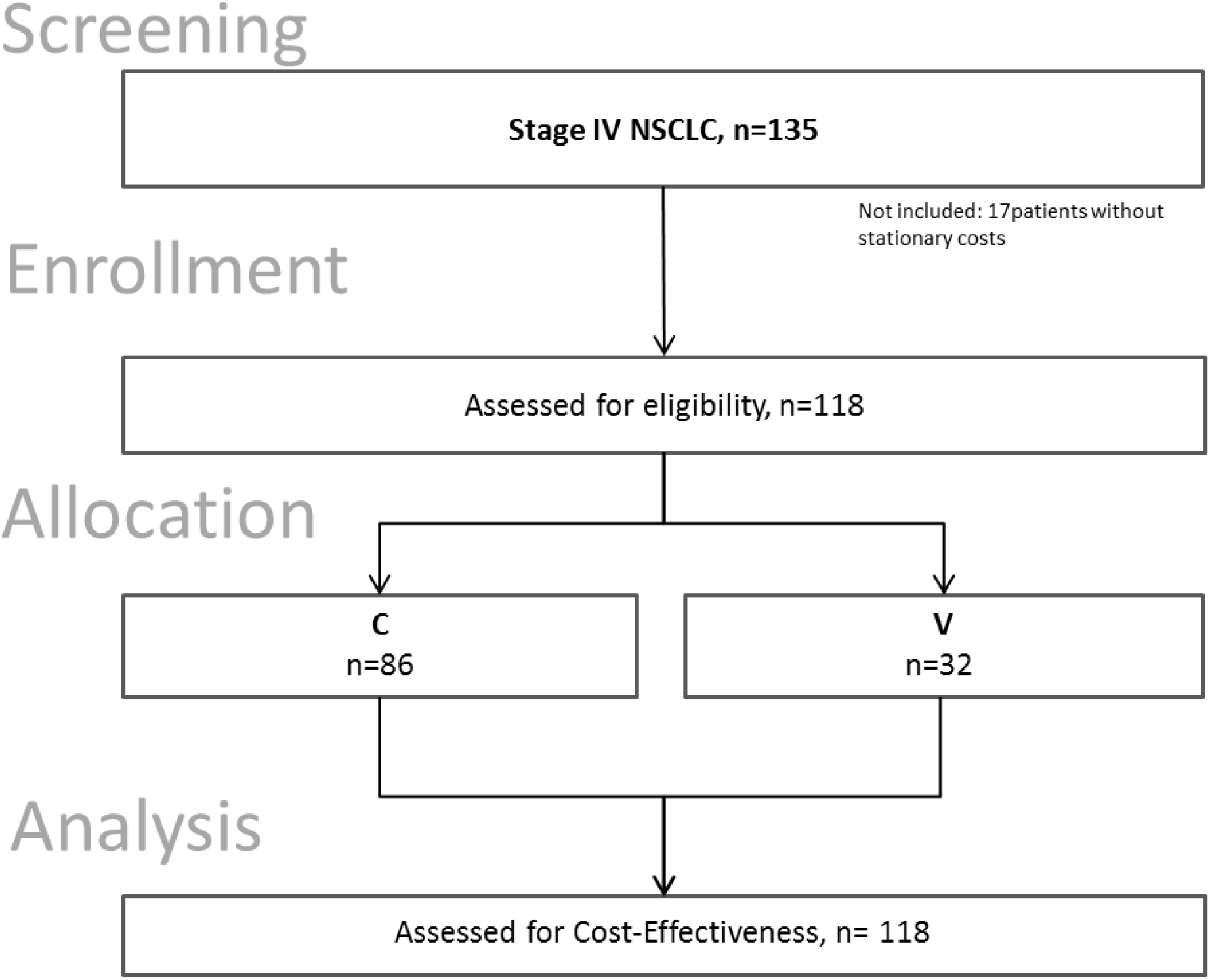
Flow chart of the study population. CE, cost-effectiveness; C, group of patients receiving chemotherapy only, V, group of patients receiving chemotherapy plus add-on Viscum album L.; VA, Viscum album L., mistletoe

The mean age of the patients was 63.8 years. Patients in the V group were on average 1.3 years older than patients in the C group (V: 64.8 years, C: 63.5 years, p=0.58). 55.1% of the patients were male with slight differences between the groups (V: 58.8% male, C: 50.0% male, p=0.11). The majority of patients were diagnosed with adenocarcinoma (70.3%), followed by squamous cell carcinoma (19.5%) and large cell carcinoma (10.2%). Most baseline characteristics between both groups were balanced, see Tab 1.

A minority of the patients (8.5%) received cancer-directed surgery and one third (22.9%) received radiation, see Tab 1. All patients that were included in the study received chemotherapy with the majority of patients receiving platinum-based chemotherapy (C: 97.7%, V: 100%) mostly in combination with pemetrex(ed) (C: 53.5%, V: 59.4%) or vinorelbine (C: 37.2%, V: 21.9%), see Tab 2. With the exception that the C-group received significantly more vinorelbine than the V-group, the therapeutic regimens between both groups were balanced.

**Tab 2:** Composition of chemotherapeutic treatment. Oncological treatment of patients per group, n=number of patients, total number of patients per group do not necessarily add to 100% as patients may have received various combinations of chemotherapy, n=number of patients, %, percent

**Tab 2. Composition of chemotherapeutic treatment**
|  | C (n=86)<br>n (%) | V (n=32)<br>n (%) | p-value |
| --- | --- | --- | --- |
| chemotherapy | 86 (100) | 32 (100) | -- |
| platinum - based | 84 (97.7) | 32 (100) | 0.39 |
| pemetrex(ed) | 46 (53.5) | 19 (59.4) | 0.57 |
| gemcitabine | 0 | 2 (6.3) | 0.13 |
| vinorelbine | 32 (37.2) | 7 (21.9) | 0.02 |
| paclitaxel | 3 (3.5) | 0 | 0.12 |

32 patients (27.1%) of the cohort received supportive *Viscum album* L. extracts in addition to chemotherapy. While the majority of these patients applied the extracts subcutaneously (100%), twenty one patients (65.6%) additionally received intravenous and two patients (6.3%) received intratumoural applications. With respect to the VA host trees most patients received add-on VA from the fir tree (81.3% VA fraxini), followed by VA from the pine tree (43.8% VA pini), and from the oak tree (37.5% VA quercus), see Tab 3.

**Tab 3:** Composition of add-on VA treatment in the V-groupy. Number of V-group patients (n=32) exposed to various add-on mistletoe (VA) extracts, pooled per host tree. n=number of patients, total number of patients per VA remedy do not necessarily add to 100% as patients may have received various combinations of VA treatment, n=number of patients, %, percent; s.c., subcutaneous; i.v., intravenous; i.t., intratumoural; ^1)^ reference for s.c. application; ^2)^ reference for i.v. application; ^3)^ reference for i.t. application

**Tab 3. Composition of add-on VA treatment in the V-group**
|  | Total n=32 (100)<br>n (%) | s.c. n=32 (100)<sup>1)</sup><br>n (%) | i.v. n=21 (100)<sup>2)</sup><br>n (%) | i.t. n=2 (100)<sup>3)</sup><br>n (%) |
| --- | --- | --- | --- | --- |
| Viscum abietis (spruce) | 1 (3.1) | 1 (3.1) | -- | -- |
| Viscum fraxini (fir tree) | 26 (81.3) | 17 (53.1) | 7 (33.3) | 2 (100) |
| Viscum mali (apple) | 5 (15.6) | 5 (15.6) | -- | -- |
| Viscum pini (pine) | 14 (43.8) | 1 (2.9) | 13 (61.9) | -- |
| Viscum quercus (oak) | 12 (37.5) | 11 (34.4) | 1 (4.8) | -- |

### Overall Survival

When patients from the V-group were compared to patients from the C-group, the multivariable cox regression analysis adjusting for age, gender, BMI, radiation, cancer-directed surgery, smoker status and add-on VA (stratifying for histology and chemotherapy) revealed an adjusted hazard risk (aHR) of add-on VA treatment of 0.37, standard error (se) 0.344, p=0.01) indicating that the hazard of dying was reduced by 63%, data not shown. Male gender increased the hazard of dying by factor 2 (aHR=1.99, se 0.232, p=0.03). Furthermore, the direction of impact on hazard of death was negative (i.e. positive for OS) for radiation (aHR=0.86) and surgery (aHR=0.39) but not significant, data not shown.

The age-adjusted mean OS for the C-group was 13.4 months and for the V-group 19.1 months, indicating that patients from the V-group lived 5.7 months longer than patients from the C-group, see Tab 4. As no death occurred in both groups before start of treatment and as time until treatment was not significantly different between both groups (p=0.69) a time-dependent bias could be precluded. The mean time from diagnosis until treatment was 32.4 days in the C-group and 37.9 days in the V-group.

### Cost Outcomes

The cost estimates for both treatment groups are also shown in Tab 4. Adjusted mean hospital’s total costs were €16,299.98 for the C-group and €17,992.26 for the V-group. Highest hospital’s expenditures per patient were documented for normal ward, radiology, endoscopic diagnostics & therapy, ICU, laboratories and diagnostics with no relevant differences between the groups, see figure 2. Further analyses revealed that, as a consequence of longer survival, patients from the V-group had on average 1.7 additional hospital stays since first diagnosis than patients from the C-group, (p>0.01), data not shown. Mean hospital visits in the C- and in the V-group were 3.8 (standard deviation, SD ±2.2) and 5.5 (SD ±3.1) visits, respectively. While patients from the C-group were on average 11.6 (±8.0) days in the hospital, patients from the V-group were on average 2.4 days shorter in the hospital with 9.2 (±4.1) days, however the difference was not significant (p=0.11).

**Tab 4:** Cost- and cost-effectiveness analyses. Cost and CEA analyses for stage IV bronchial cancer; ^1^adjusted for age, gender, BMI, histology, smoker status, surgery, radiation and chemotherapy

**Tab 4. Cost- and cost-effectiveness analyses**
|  | C | V |
| --- | --- | --- |
| adjusted average total costs<sup>1</sup><br>€, (95%CI) | 16,288.98<br>(13,833.86-18,744.10) | 17,992.26<br>(13,658.26-22,326.26) |
| adjusted restricted mean OS<sup>1</sup><br>months, (95%CI) | 13.4 (10.4-16.4) | 19.1 (12.4-25.8) |
| adjusted average cost per adjusted mean months OS<br>€/months | 1,215.60 | 942.00 |
| stationary costs per gained life month (ICER) for V vs. C<br>€/months |  | 298.82 |
| **stationary costs per gained life year (ICER) for V vs. C**<br>**€/year** |  | **3,585.84** |

**Fig 2:**
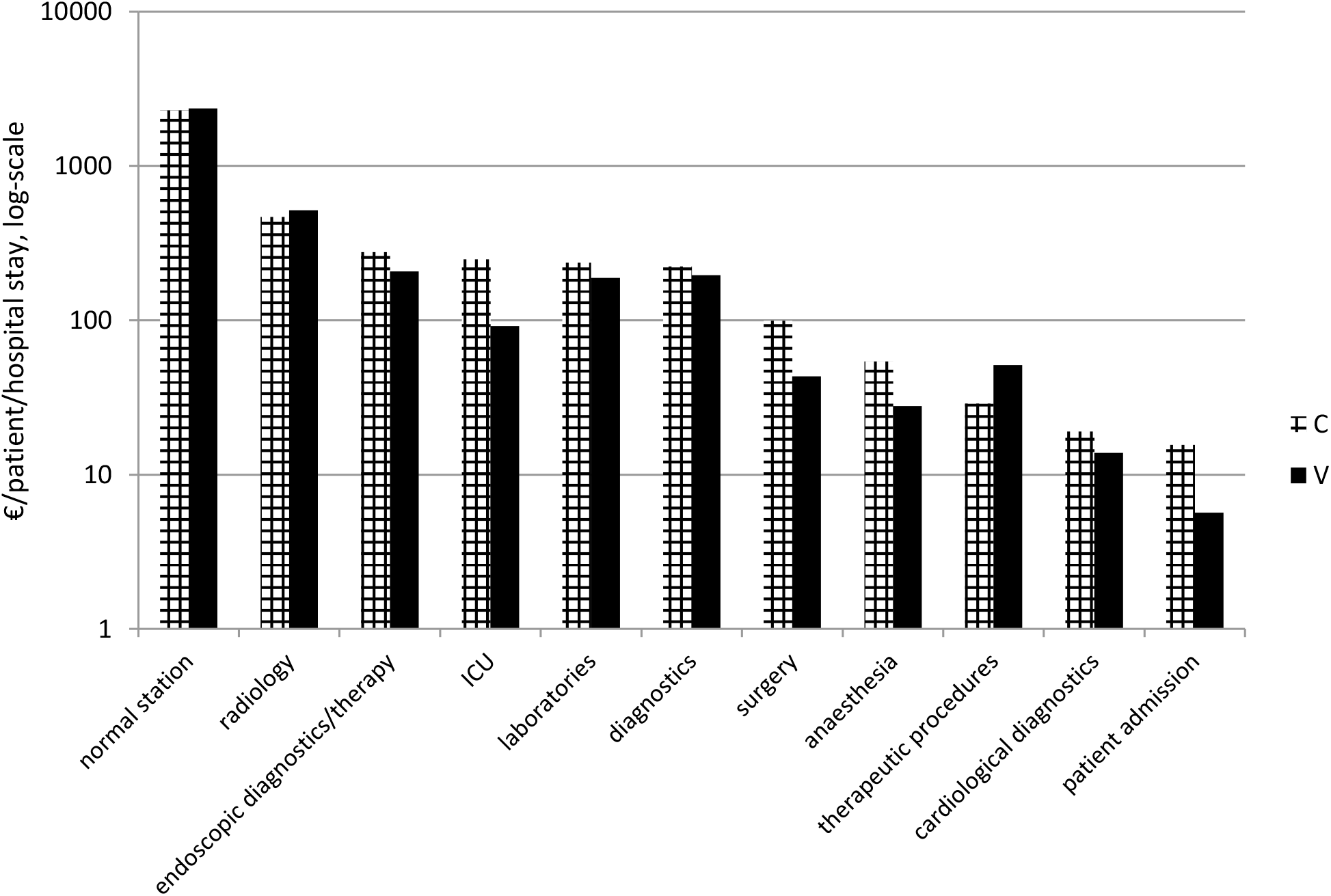
Mean hospital’s costs per patient per hospital stay according to treatment group C and V, logarithmical y-scale; €, Euro

Multivariable regression analysis revealed that male gender (estimate -6,570.47, se 2,270.73, p=0.005), the histology of a squamous cell carcinoma (estimate -5,575.06, se 2,756.42, p=0.046) and V-patients that have died (estimate -10,356.58, se 4,893.26, p=0.03) were significantly associated with less hospital expenditures, while the overall incidence of death of patients was significantly associated with increased hospital costs (estimate 6,368.96, se 2,674.52, p=0.02), data not shown.

### Cost-Effectiveness

The survival analyses demonstrated that patients in the V-group showed longer survival than those receiving standard of care (a 5.7 month longer adjusted overall mean survival). Cost analyses have shown that this longer survival was associated with additional total hospital costs (Tab 4). Division of adjusted average costs by adjusted mean overall survival revealed for C-treatment mean costs of €1,215.60 per mean month OS and for the V-treatment €942.00 per mean month OS. Compared to C, patients with V-treatment had relevant hospital’s savings of €273.6 per mean OS. We calculated an ICER of €298.82 per additional month OS and of €3,585.84 per additional year OS resembling the costs for the improvement of one OS year gained with the V-treatment compared to the C-treatment. For the outcome mean costs per patient from the hospital perspective in combination with mean months OS a probabilistic sensitivity analysis was performed. Fig 3 shows a scatter plot of all replicated results for bootstrap sensitivity analyses. Most single dots are located in the upper right-hand quadrant indicating a probability of 73.6% and confirming the robustness of the CEA outcome. Nevertheless, a relevant proportion of single dots were located in the lower right-hand quadrant pointing to a 26.4% probability of hospital’s cost savings.

**Fig 3:**
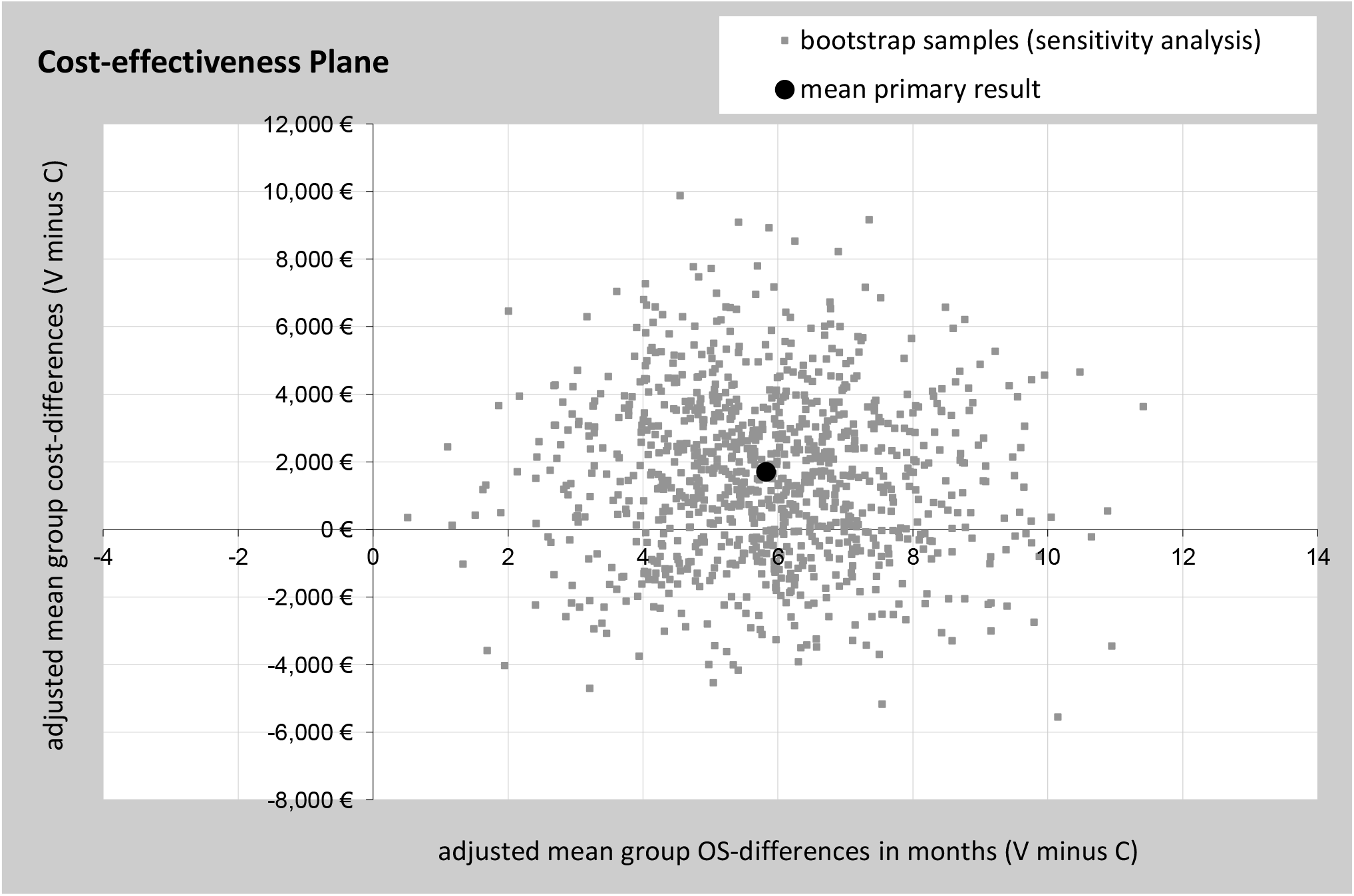
Sensitivity analyses of the outcome cost per months OS from the hospital’s perspective. Incremental cost-effectiveness plane showing random 1000-fold resampled estimates (bootstrap analysis) of incremental costs and benefits (age-adjusted mean cost per patient in combination with age-adjusted mean months OS) of using V-treatment for stage IV NSCLC patients versus C-treatment

## Discussion

Lung cancer healthcare costs generally are on the rise due to the high costs of rapidly progressing molecular-based personalized therapies. Cost and cost-effectiveness investigations of integrative oncology concepts are increasingly becoming an additional tool for hospital systems, health administrations and decision makers. This is due to the increase in cancer burden (population growth, aging, lifestyle changes) and increasing patient’s demand for active participation in the decision to receive treatments, which contribute to maintaining HRQl and tolerance of standard oncology therapies. The objective of the study was the cost-effectiveness analysis of combinatorial CTx plus VA compared to CTx alone in stage IV NSCLC patients from the hospital’s perspective.

Our analysis showed that patients treated with a combination of CTx and VA received a cost-effective therapy with savings per overall survival achieved compared to CTx. We demonstrated that patients receiving combinatorial treatment had a longer mean OS and a significant association with a reduced risk of death compared to patient’s only receiving CTx alongside with the results of a recent study in patients with stage IV NSCLC [19]. These results are consistent with other published data on the clinical effects of combinatorial VA therapy in cancer patients [20-23]. The relevant negative association of the male gender with survival was observed in our patients and correlated with other published data [24].

Cost analysis showed that V treatment resulted in higher hospital spending, which can be explained by the higher number of hospital stays due to longer survival. Nevertheless, patients with combinatorial treatment (V-group) had a shorter length of hospital stay compared to patients treated with CTx only (C-group). A shorter length of stay means lower hospital costs for treatment, services and medication. However, according to balanced co-morbidities in both groups V-patients were not healthier than C-patients. Thus, the longer hospital stay of C-patients stays remains unclear. Male gender was significantly associated with reduced hospital costs, and it is assumed that this is caused by the significantly shorter survival versus female patients. This is consistent with the observation of another lung cancer cost study [25], which shows average hospital care costs (male: €446, female: €615, p = 0.048), services (male: €946, female: €1.395, p = 0.049) and medical care (male: €117, female: €214, p = 0.006) being statistically lower in men because of their shorter survival (p = 0.045) [26] than in female lung cancer patients. Further, our cost analysis revealed that the incidence of death was associated with higher hospital cost burden. This is in line with the results of other studies published indicating that end of life is one of the most expensive periods of cancer treatment [27]. Calculations of an actual in-depth cost study on 230 patients with the majority of NSCLC (78%) in an advanced stage revealed that the key drivers of the last six month of life were chemotherapy (39%), concomitant medication (14%) and cost of hospitalization (14%) [28].

Our cost-effectiveness analysis reveal, that the combinatorial therapy was not inferior against the treatment standard, as it resulted in relevant hospital savings per gained survival year compared to the control. The calculation of the incremental cost-effectiveness ratio (ICER) yielded a value of € 3,585.84 per gained surviving year. On the assumption that combinatorial treatment is more effective and at least equivalent in maintaining quality of life, this ICER-value appears low and affordable in modern oncological settings. Despite ICERs being common references in health economics, their values are difficult to compare between studies depending on the perspective (e.g. hospital, society etc.), the end point (e.g. life years, QALYs etc.), the respective costs (e.g. direct and/or indirect costs; inpatient and/or outpatient costs etc.) and country to list just a few factors. In the field of pulmonary oncology, there are published ICERs on first-line tyrosine kinase inhibitor treatment in advanced NSCLC ranging from $78,514 (converted into Euro: €71,250) to $100,120 (converted into €90,857) per gained surviving year compared to cisplatin-pemetrexed regimen in advanced NSCLC patients [29]. As to second-line PD-1 immunotherapies for these tumors the range is between $114,303 and $249,169 (converted into €103,728-€226,116) per life year gained versus docetaxel [29]. Thus, the comparability to our data is limited because the ICERs in our data relate to real-world German hospital costs. Nevertheless, against the background of rapidly rising lung cancer healthcare costs, cost effectiveness data from real-world research will make a valuable contribution in the future and will become increasingly relevant. Among the limitations of the present analysis are the monocentric, uncontrolled and observational nature of the study and the limited transferability of cost and CE data to other countries. The strength of this study lies in its pragmatic design, the integration of real-world daily care data under typical hospital conditions and the sensitivity analysis of the model confirming the results to be robust. The external validity of our data may be of clinical weight, as the characteristics of the patient, the type of treatment and the outcome are comparable to published data. In the future, randomized, multi-centre, prospective cost-benefit analyses including outpatient cost data should limit the potential bias and create a more comprehensive cost spectrum for IO in lung cancer.

To our knowledge, this is the first study to directly analyse the cost and cost effectiveness of chemotherapy plus VA therapy compared to chemotherapy alone in stage IV NSCLC patients based on real world patient data and hospital costs in Germany.

## Conclusions

Cost and cost effectiveness analyses of IO in lung cancer are rare. This first comprehensive assessment of inpatient costs and cost-effectiveness of IO concepts in stage IV NSCLC suggests that the combined use of chemotherapy and VA is clinically effective and comparably cost-effective to chemotherapy alone in our analysed patient sample. For reassessment of our results, randomized and prospective health economic studies are essential.

## Data Availability

All relevant data are within the manuscript and its supporting information files.

## Availability of data and material

All relevant data are within the manuscript and its supporting information files.

